# Early left ventricular systolic function is a more sensitive predictor of adverse events after heart transplant

**DOI:** 10.1101/2023.03.21.23287555

**Authors:** Zhenxing Sun, Yu Cai, Yujia Yang, Lei Huang, Yuji Xie, Shuangshuang Zhu, Chun Wu, Wei Sun, Ziming Zhang, Yuman Li, Jing Wang, Lingyun Fang, Yali Yang, Qing Lv, Nianguo Dong, Li Zhang, Haotian Gu, Mingxing Xie

## Abstract

**Background:** First-phase ejection fraction (EF1) is a novel measure of early systolic function. This study was to investigate the prognostic value of EF1 in heart transplant recipients.

**Methods:** Heart transplant recipients were prospectively recruited at the Union Hospital, Wuhan, China between January 2015 and December 2019. All patients underwent clinical examination, biochemistry measures [brain natriuretic peptide (BNP) and creatinine] and transthoracic echocardiography. The primary endpoint was a combined event of all-cause mortality and graft rejection.

**Results:** In 277 patients (aged 48.6±12.5 years) followed for a median of 38.7 (interquartile range: 18.3) months, there were 35 (12.6%) patients had adverse events including 20 deaths and 15 rejections. EF1 was negatively associated with BNP (β=-0.220, p<0.001) and was significantly lower in patients with events compared to those without. EF1 had the largest area under the curve in ROC analysis compared to other measures. An optimal cut-off value of 25.8% for EF1 had a sensitivity of 96.3% and a specificity of 97.1% for prediction of events. EF1 was the most powerful predictor of events with hazard ratio per 1% change in EF1: 0.628 (95%CI: 0.555-0.710, p<0.001) after adjustment for left ventricular ejection fraction and global longitudinal strain.

**Conclusions:** Early left ventricular systolic function as measured by EF1 is a powerful predictor of adverse outcomes after heart transplant. EF1 may be useful in risk stratification and management of heart transplant recipients.

## Introduction

Heart transplant (HT) is the treatment of choice for patients with advanced (end-stage) heart failure. Over the last decades, heart transplants reported to the registry of the International Society for Heart and Lung Transplantation (ISHLT) have been steadily increasing (1). Although the advance in immunosuppression therapy has increased the survival rate after HT (1), cardiac allograft vasculopathy (CAV), acute cellular rejection (ACR) and antibody-mediated rejection (AMR) remain as major causes of cardiac complications (2-4).

Therefore, close monitoring of graft function is essential. Transthoracic echocardiography as a most widely used cardiac imaging modality has been recommended for routine surveillance of graft function by the American Society of Echocardiography (ASE) (5) and the European Association of Cardiovascular Imaging (EACVI) (6). However, recent studies indicated that left ventricular ejection fraction (EF) and diastolic functional measures may not be the most optimal markers for detection of early graft failure (7,8). Left ventricular longitudinal strain (LVGLS) has been shown to have prognostic value of adverse cardiac events (9,10). However, LVGLS is still limited by the lack of standardization between various vendor platforms with lack of uniform established reference values. There is an ongoing search for more sensitive imaging biomarkers of early rejection, which can (partially) supplant regular endomyocardial screening biopsies and their attendant risks.

First-phase ejection fraction (EF1), a novel but simple measure of early systolic function, is a sensitive marker of early systolic impairment in patients with aortic stenosis, COVID-19 and a powerful predictor of response to cardiac resynchronization therapy (11-14). The impairment of early systolic function could be related to diastolic relaxation and dysfunction through the shortening-deactivation phenomenon: shortening of the myocyte after depolarization leads to alterations in the cytosolic calcium transient and reduced contraction (15-17). Therefore, the objective of the present study was to examine the relative prognostic impact of EF1 compared to other measures of systolic function including LVEF and LVGLS, diastolic function and brain natriuretic peptide (BNP) in heart transplant recipients.

## Methods

### Patient population

A prospective observational study of the predictive value of EF1 for adverse events was undertaken in consecutive HT patients at The Union Hospital, Wuhan, China between January 2015 and December 2019. The study was approved by the Union Hospital Tongji Medical College Ethical committee and complied with the Declaration of Helsinki. All adult patients underwent clinical examination, biochemistry measures (BNP and creatinine) and transthoracic echocardiography. All patients were followed until defined endpoint or censoring date on 31st December 2021. All patients received immunosuppression regimens (including tacrolimus, and mycophenolate mofetil).

### Rejection

Endomyocardial biopsies (EMB) was performed as part of post-heart transplantation evaluation based on the standard institutional follow-up protocol. A minimum of 3 ventricular myocardial fragments (each consisting of at least 50% myocardium) were obtained and analysed by the pathology laboratory.

Cellular rejection was graded according to ISHLT grading system (18): grade 0R biopsies being classified as negative for cellular rejection, grade 1R biopsies as mild cellular rejection, and grades 2R and 3R biopsies as significant cellular rejection. Rejection in the present study was defined as Grade ≥2R cellular rejection on histopathology or as antibody-mediated rejection (18).

### Endpoint

The primary endpoint was defined as a combined event: all-cause mortality and graft rejection. Outcome data were verified from hospital records and death registry database.

### Transthoracic echocardiography and first phase ejection fraction

Transthoracic echocardiography was performed using a Philips Epiq 7C and IE Elite (Philips Healthcare, Guildford, UK) ultrasound platforms. All echocardiographic measurements were performed according to the recommendations of the American Society of Echocardiography (19). Image analysis was performed by two authors blinded to clinical information and outcomes using a Tomtec analysis package (Tomtec, Munich, Germany). Left atrial volume (LAV) was measured from apical 4 and 2 chamber views. LAV index (LAVi) were obtained from BSA. Stroke volume (SV) was calculated as the difference between end-diastolic volume (EDV) and end-systolic volume (ESV) and indexed to BSA to give stroke volume index (SVi). Tissue Doppler measures were obtained at levels of the lateral and septal mitral annulus to obtain an optimal spectral Doppler waveform. E/e’ ratio was calculated as a measure of diastolic function from the ratio of transmitral Doppler E wave velocity to the mean of basal lateral and septal tissue Doppler e’ waves (20). Left ventricular global longitudinal strain was measured from speckle tracking echocardiography from apical views.

LV volumes were measured by 2D Simpson’s method from apical views at end-diastole, time of peak aortic valve flow velocity (TPAVF) and end-systole. LVEF was calculated as the percentage change of LV volume from end-diastole to end-systole. EF1 was taken as the percentage change in LV volume from end-diastole to TPAVF, a time that approximates the time of peak ventricular contraction and expressed as a percentage of EDV (figure 1). The frame from apical views for determining LV volume at TPAVF is estimated by the measuring the total number of frames from the R wave to the end systole and multiplying this by the fraction of time from the R wave to TPAVF and from R wave to end systole.(21) Right ventricular fractional area change (RVFAC) was measured as percentage change between RV area at the end-diastole and area at the end-systole. Tricuspid annular plane systolic excursion (TAPSE) was measured using M-mode from an apical 4-chamber view.

**Figure 1:**
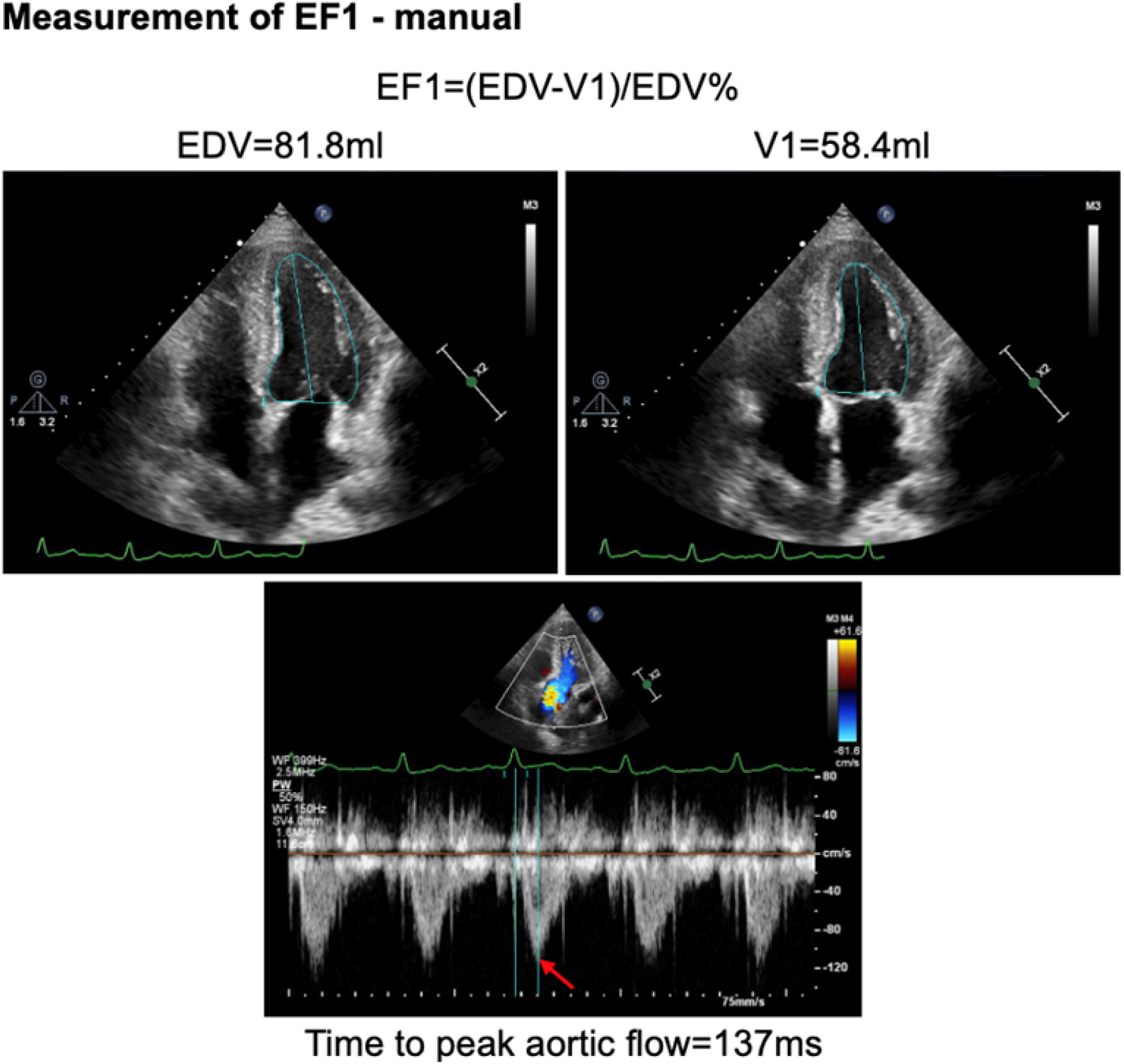
Measurement of EF1

### Repeatability and reproducibility of EF1

Intra- and inter-observer variability in measurements of EF1 was assessed in 30 randomly selected subjects by measurements repeated 2 months apart by 2 observers with the standard deviation of difference in measures on the two occasions and coefficient of variation (CV) used as a measure of variability.

### Statistical Analysis

Continuous variables were tested for normality, and those following an approximately normal distribution are presented as means±standard deviation (SD). Other variables are presented as medians and inter-quartile range. Comparisons between groups were made by analysis of variance or by χ^2^ test for categorical variables. For the prediction of events, receiver-operating characteristic (ROC) analysis was performed to examine the sensitivity and specificity of EF1 for events and the optimal cut-off value for predicting events was determined to maximise values of sensitivity and specificity. The added discriminative power of EF1 over other measures was tested using the C-statistic. Kaplan-Meier curves were used to examine cumulative event rates and the difference between groups was tested using a log rank test. Univariable Cox regression analysis was performed to identify potential predictors of events. Multivariable Cox proportional hazards models were used to test the independent value of echocardiographic and other measures for predicting future events. A two-tailed P-value of <0.05 was considered statistically significant. All statistical analyses were performed using SPSS 24 for Mac (SPSS Inc. Chicago, IL, USA).

## Results

### Characteristics of patients

300 consecutive patients underwent heart transplant were recruited of which 23 were excluded from the final analysis, due to poor image quality (n=19), lost to follow-up (n=4). Thus, a total of 277 patients (aged 48.6±12.5 years) were included in the final analysis. Echocardiography was performed at median of 11.1 (interquartile range:15.9) months after HT. After a median of 38.7 (interquartile range: 18.3) months of follow-up, 35/277 (12.6%) patients reached endpoints including 20 deaths and15 rejections. Event-free survival was 94.6%, 92.4% and 88.8% at 12, 24 and 36 months respectively.

### Clinical, laboratory and echocardiographic data

EF1 was negatively associated with BNP with (β=-0.161, p=0.003) or without (β=-0.220, p<0.001) adjustment for recipient age, gender, LAVi, e’, LVEF, LVGLS, and TAPSE.

When comparing patients with events to those without events, there was no significant difference in donor age, recipient age, recipient gender, BMI, heart rate, systolic and diastolic blood pressures, history of hypertension and diabetes. Patients with events had significantly higher creatinine and BNP compared to those without events. Patients with events had worse LV systolic function (as measured by LVEF, EF1 and LVGLS), diastolic relaxation (as measured by e’) and increased LAVi, but no difference in RV systolic function (table 1).

**Table 1.** Baseline characteristics in patients with and without events.

|  | **Events (n=35)** | **No Events (n=242)** | **P** |
| --- | --- | --- | --- |
| **Recipient Age (years)** | 50.0±12.1 | 48.5±12.6 | 0.520 |
| **Donor Age (years)** | 46.0±13.7 | 47.5±12.3 | 0.527 |
| **Recipient Male Sex (%)** | 28 (80.0) | 198 (81.8) | 0.816 |
| **Body Mass Index (kg/m<sup>2</sup>)** | 22.5±4.5 | 23.2±3.5 | 0.339 |
| **Heart rate (bpm)** | 91±9 | 91±10 | 0.907 |
| **Systolic Blood Pressure (mmHg)** | 119±20 | 119±11 | 0.750 |
| **Diastolic Blood Pressure (mmHg)** | 80±11 | 79±10 | 0.544 |
| **Reason for transplant** |  |  | 0.804 |
| **Dilated Cardiomyopathy (%)** | 17 (48.6) | 143 (59.1) |  |
| **Hypertrophic Cardiomyopathy (%)** | 1 (2.9) | 9 (3.7) |  |
| **Ischemic Heart Disease (%)** | 7 (20) | 38 (15.7) |  |
| **Heart Valve Disease (%)** | 3 (8.6) | 21 (8.7) |  |
| **Adult Congenital Heart Disease (%)** | 2 (5.7) | 11 (4.5) |  |
| **Other (%)** | 5 (14.3) | 20 (8.3) |  |
| **Co-morbidities** |  |  |  |
| **Hypertension** | 21 (60.0) | 111 (45.9) | 0.148 |
| **Diabetic Mellitus** | 24 (68.9) | 129 (53.3) | 0.103 |
| **Medications** |  |  |  |
| **Tacrolimus** | 21 (60.0) | 134 (55.4) | 0.606 |
| **Mycophenolate mofetil** | 20 (57.1) | 130 (53.7) | 0.538 |
| **ACE/ARB** | 6 (17.1) | 35 (14.5) | 0.618 |
| **Clopidogrel** | 4 (11.4) | 14 (5.8) | 0.260 |
| **CCB** | 16 (45.7) | 93 (38.4) | 0.464 |
| **Statins** | 22 (62.9) | 142 (58.7) | 0.854 |
| **Aspirin** | 7 (20) | 81 (33.5) | 0.123 |
| **Biochemistry** |  |  |  |
| **Creatinine (umol/l)** | 121.5±62.5 | 105.8±33.7 | **0.025** |
| **BNP (pg/ml)** | 204.3±275.2 | 112.2±92.1 | **<0.001** |
| **Echocardiography** |  |  |  |
| **LV End Diastolic Volume (ml)** | 90.1±24.3 | 88.7±18.9 | 0.689 |
| **Stroke Volume index (ml/m<sup>2</sup>)** | 31.9±10.3 | 32.3±6.6 | 0.760 |
| **Left Atrial Volume index (ml/m<sup>2</sup>)** | 49.3±15.5 | 42.4±11.7 | **0.002** |
| **LV Ejection Fraction (%)** | 59.2±4.3 | 62.1±3.8 | **<0.001** |
| **EF1 (%)** | 21.2±2.6 | 30.4±3.1 | **<0.001** |
| **LV Global Longitudinal Strain (%)** | -16.1±2.2 | -19.5±2.1 | **<0.001** |
| **S wave (cm/s)** | 8.5±1.7 | 8.8±1.4 | 0.229 |
| **e' wave (cm/s)** | 11.3±2.8 | 12.6±3.2 | **0.024** |
| **E/e'** | 7.6±3.2 | 7.0±2.7 | 0.203 |
| **TAPSE (mm)** | 14.0±4.0 | 15.4±6.5 | 0.202 |
| **RV FAC (%)** | 43.6±7.8 | 46.2±15.7 | 0.330 |
| **TPAVF (ms)** | 128.1±16.6 | 125.5±19.4 | 0.450 |
| **TPAVF/Ejection Time** | 0.45±0.05 | 0.44±0.06 | 0.461 |

ACE: angiotensin converting enzyme; ARB: angiotensin receptor blockers; CCB: calcium channel blockers; BNP: brain natriuretic peptide; LV: left ventricle; EF1: first-phase ejection fraction; S: tissue Doppler mitral annulus systolic motion; e': tissue Doppler mitral annulus early diastolic motion; E/e': early mitral filling / tissue Doppler mitral annulus early diastolic motion; TAPSE: tricuspid annular plane systolic excursion; FAC: fractional area change; TPAVF: time to peak aortic valve flow velocity.

Intra- and inter-observer coefficients of variation for EF1 were 2.1% and 3.2%, respectively. Bland-Altman plots for inter and intra-observer variability are shown in figure 2.

**Figure 2:**
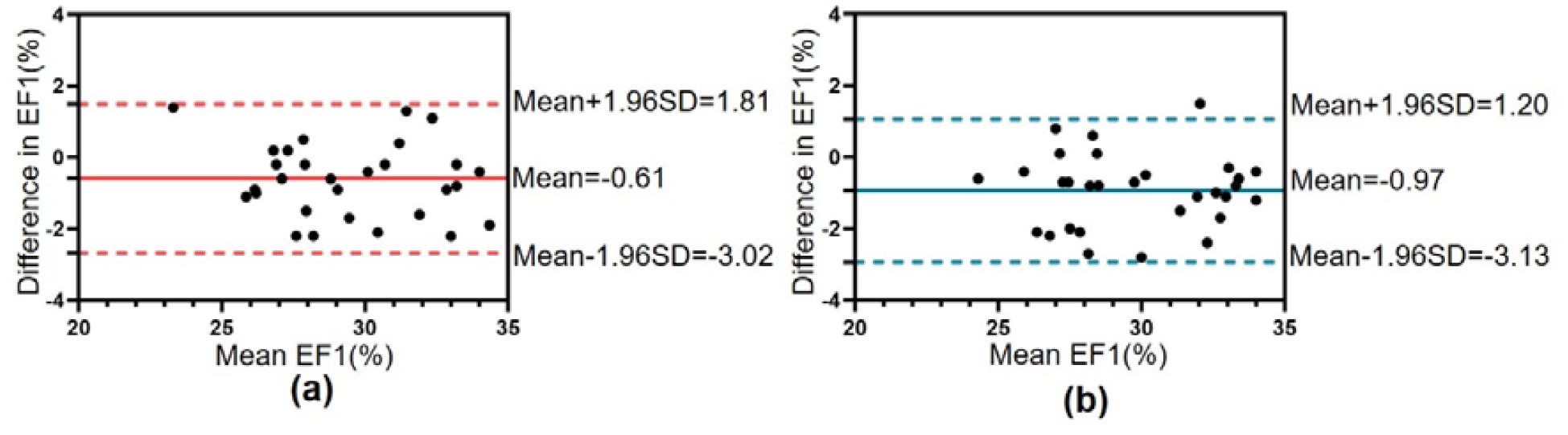
Bland-Altman plots of intra (a) and inter (b) observer viability of EF1

### Prediction of events by EF1 and other measures

Prediction of events by EF1 and other measures by univariate and multivariate Cox regression analysis is shown in table 2. EF1 was the most consistently and strongly predictive of events in univariate and each of multivariate models. Other measures significantly predicting events in univariate analysis included: BNP, Creatinine, LAVi, LVEF, LVGLS, e’, and TAPSE (table 2). In multivariable model 1, including significant predictors in the univariable analysis with a p<0.001 (LVEF, LVGLS and EF1), EF1 was the only measure strongly associated with events [hazard ratio (HR) per 1% change in EF1: 0.628, 95% confidence interval (CI): 0.555-0.710, p<0.001]. Similar HR for EF1 was observed (0.623, 95%CI: 0.548-0.708, p<0.001) when all significant predictors (p<0.05) in the univariable analysis were included (multivariable model 2). ROC curve analyses of EF1, BNP, LAVi, LVEF, LVGLS and TAPSE for predicting events are shown in figure 3. The area under the curve (AUC) was the largest for EF1 (0.987, p<0.001), followed by LVGLS, LVEF, LAVi, BNP and TAPSE. An optimal cut-off value of 25.8% for EF1 had a sensitivity of 96.3% and a specificity of 97.1% for prediction of events. The C-statistic index for a logistic model (including variables from Cox regression multivariable model 2) increased significantly by adding EF1 (0.897 to 0.989: change in the c-statistics 0.092, p<0.05).

**Table 2.** Univariable and multivariable analysis of predictors of events.

|  | **HR** | **CI (95%)** | **P value** |
| --- | --- | --- | --- |
| **Univariable** |  |  |  |
| Donor Age | 0.991 | 0.965 – 1.017 | 0.481 |
| Recipient Age | 1.009 | 0.982 – 1.036 | 0.514 |
| Recipient Gender | 0.906 | 0.396 – 2.076 | 0.816 |
| Body Mass Index | 0.950 | 0.865 – 1.044 | 0.289 |
| Heart Rate | 1.002 | 0.969 – 1.036 | 0.915 |
| Systolic Blood Pressure | 1.003 | 0.976 – 1.032 | 0.823 |
| Hypertension | 0.591 | 0.300 – 1.161 | 0.127 |
| Diabetes | 0.532 | 0.260 – 1.085 | 0.083 |
| Adult Congenital Heart Disease | 0.830 | 0.199 – 3.461 | 0.798 |
| ln BNP | 1.808 | 1.176 – 2.778 | **0.007** |
| ln Creatinine | 3.012 | 1.058 – 8.578 | **0.039** |
| Stroke Volume index | 0.996 | 0.949 – 1.045 | 0.868 |
| Left Atrial Volume index | 1.035 | 1.012 – 1.058 | **0.002** |
| LV End-diastolic Volume | 1.004 | 0.988 – 1.021 | 0.596 |
| LV Ejection Fraction | 0.842 | 0.777 – 0.912 | **<0.001** |
| LV Global Longitudinal Strain | 1.634 | 1.455 – 1.833 | **<0.001** |
| e' | 0.879 | 0.790 – 0.978 | **0.018** |
| E/e' | 1.078 | 0.973 – 1.195 | 0.153 |
| TPAVF | 1.007 | 0.990 – 1.025 | 0.426 |
| EF1 | 0.625 | 0.565 – 0.691 | **<0.001** |
| TAPSE | 0.884 | 0.800 – 0.977 | **0.016** |
| RV FAC | 0.954 | 0.905 – 1.007 | 0.086 |
| **Multivariable Model 1** |  |  |  |
| Ejection Fraction | 1.068 | 0.975 – 1.170 | 0.158 |
| Global Longitudinal Strain | 1.091 | 0.891 – 1.336 | 0.397 |
| EF1 | 0.628 | 0.555 – 0.710 | **<0.001** |
| **Multivariable Model 2** |  |  |  |
| Ln BNP | 0.835 | 0.554 – 1.257 | 0.387 |
| Ln Creatinine | 1.591 | 0.447 – 5.662 | 0.473 |
| Left Atrial Volume Index | 1.007 | 0.981 – 1.035 | 0.585 |
| Ejection Fraction | 1.052 | 0.955 – 1.158 | 0.305 |
| Global Longitudinal Strain | 1.055 | 0.864 – 1.287 | 0.775 |
| e' | 0.900 | 0.788 – 1.027 | 0.118 |
| TAPSE | 0.892 | 0.803 – 0.991 | **0.034** |
| EF1 | 0.623 | 0.548 – 0.708 | **<0.001** |

ln BNP: natural log brain natriuretic peptide; e': tissue Doppler mitral annulus early diastolic motion; E/e': early mitral filling / tissue Doppler mitral annulus early diastolic motion; TPAVF: time to peak aortic valve flow velocity; EF1: first-phase ejection fraction; TAPSE: tricuspid annular plane systolic excursion; FAC: fractional area change.

**Figure 3:**
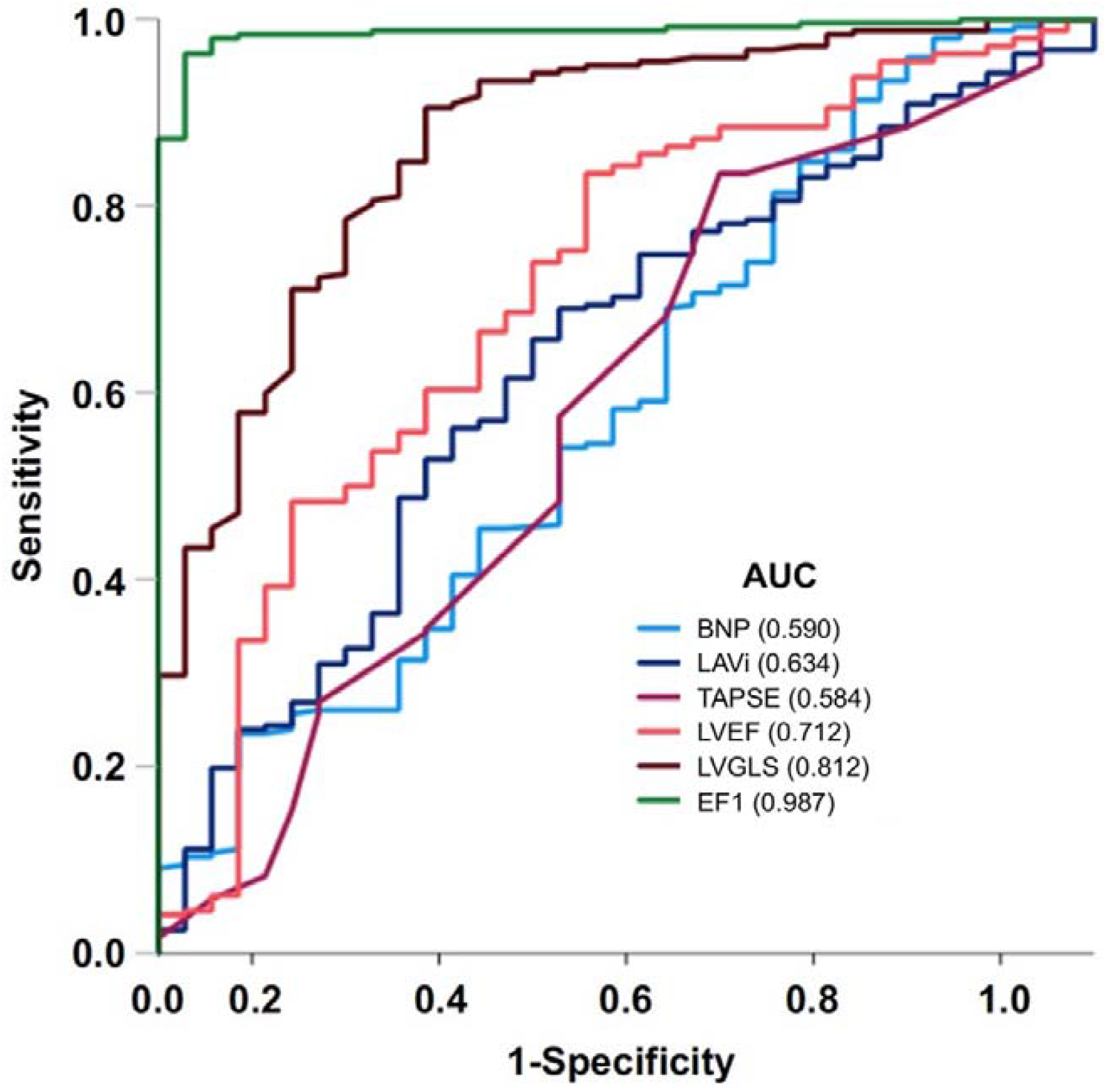
ROC analysis

Kaplan-Meier analysis showed that EF1 was a strong predictor of events (figure 4). When EF1 was less than 25.8%, 34 patients had events compared to only 1 in those with an EF1≥25.8%.

**Figure 4:**
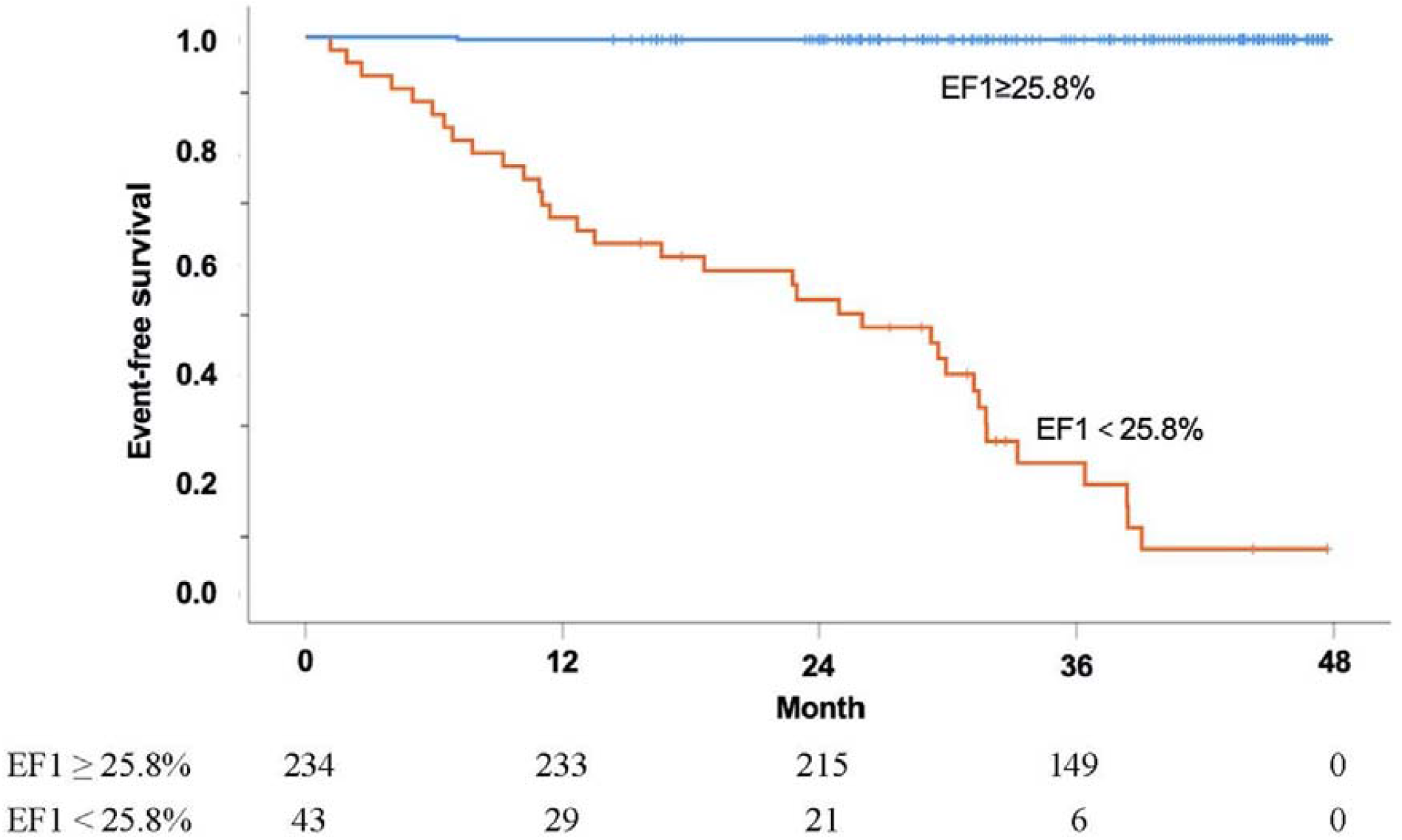
Kaplan-Meier curve for event-free survival according EF1 (cut off value 25.8%)

## Discussion

The main finding of the present study is a strong association between EF1, a measure of early systolic function and adverse outcomes in patients post HT. The finding is robust being seen irrespective of adjustment for biochemistry biomarkers and other echocardiographic measures of LV and RV function. More importantly, EF1 was superior to LVGLS and other measures of LV systolic function or RV systolic function. There is an incremental prognostic value for EF1 over other clinical and functional predictors. These findings suggest an important role for EF1, a simple but robust measure of early LV systolic function in the risk stratification of HT recipients.

Other prognostic markers in the present study included BNP, LV systolic functional measures (including LVEF and LVGLS), LV diastolic measure (e’), and TAPSE. These findings are consistent with previous studies (7-10,22-24).

### Rationale and associations of LV EF1 in heart transplant recipients

EF1 is a novel measure of early LV systolic function. Although overall LVEF and LVGLS were marginally lower in patients with events, EF1 was approx. 30% lower in patients with events compared those without. This may be because early LV systolic function, is more sensitive to compromise in cardiac contractility caused by acute or chronic injury with sarcomeric mechanotransduction maintaining overall contraction/shortening (such as overall LVEF and GLS) at the expense of slower but sustained contraction during systole (25). Less severe damage may impair the ability of the muscle to contract in early systole. Our previous studies have suggested that EF1 is associated with elevated LV myocardial wall stress in hypertensive patients (14) and myocardial fibrosis in patients with aortic stenosis (13). Cardiac allograft vasculopathy is known to affect both epicardial vessels and microvascular function which leads to impairment of early systolic function (4). Elevated myocardial wall stress induced by increased left ventricular filling pressure is common in long-term heart transplant recipients (26,27). Our finding of association between BNP and EF1 confirms that myocardial wall stress abnormalities may lead to reduced EF1 and sustained systolic contraction (13,14).

### Prognosis of EF1

EF1 is an independent predictor of adverse events in our cohort of HT recipients. Previous studies have demonstrated prognostic value of LVGLS in short- and long-term HT patients (9,23), which are consistent with our findings of LVGLS being a significant predictor of events in univariate Cox-regression analysis. In contrast to previous studies, we showed that EF1 is the only LV systolic measure remained as a significant predictor in both multivariate models. It is known that LVGLS is associated with perfusion defect, elevated LV wall stress, myocardial fibrosis and myocardial edema which are often seen in HT patients. However, LV GLS is a measure of myocardial fibre shortening through the entire systole which shares the same limitation as LVEF. The possible explanation is that the reduction of early systolic function characterised by an impaired EF1 is associated with subendocardial myocardial fibrosis and elevated myocardial wall stress.

### Clinical Perspective

EF1 is a novel but simple marker of early left ventricular systolic function, which can be easily measured from standard clinical echocardiographic examinations, and it is reproducible and vendor independent compared to GLS. EF1 is more sensitive in detecting early systolic impairment and has better predictive power than EF and GLS, therefore, EF1 has the potential to be incorporated into routine assessments in heart transplant recipients without extra training and financial burden.

### Limitations

Our study was subject to several important limitations. The number of patients was relatively small, and we cannot extrapolate our results outside of the inclusion criteria for the present study. A multi-centre study of EF1 is required to confirm its predictive value in routine clinical use. The observation of a relationship between EF1 and BNP supports an interaction between myocardial wall stress and early systolic function, but we cannot be certain of the direction of causality. Although reproducible, EF1 was measured from 2D bi-plane Simpson’s method. Such measurements are limited by geometrical assumptions and measurements made in a routine clinic may exhibit more variability. It is well recognised that automated 3D analysis and Cardiac Magnetic Resonance (CMR) provide more reliable and reproducible measurements of LV volumes and function. Therefore, EF1 measured by 3D echo or CMR would be the preferred method for future studies. The number of events following HT was low and further studies will be required to define the predictive value of EF1 for predicting clinical outcomes in these patients.

## Conclusion

In conclusion, early systolic function as captured by first-phase ejection fraction is a powerful predictor of adverse clinical outcomes. If these findings are confirmed in larger multi-centre studies and randomised controlled trials, EF1 may be useful in risk stratification and guide management of heart transplant recipients.

## Data Availability

The data that support the findings of this study are available on request from the corresponding author.

## Abbreviations

BSA: Body surface area
BNP: Brain natriuretic peptide
CV: coefficient of variation
EF1: First-phase ejection fraction
EF: Ejection fraction
EDV: End-diastolic volume
ESV: End-systole volume
HT: Heart transplant Stroke
LAV: Left atrial volume
LAVi: Left atrial volume index
LV: Left ventricular
LVGLS: Left ventricular global longitudinal strain
RV: Right ventricular
RVFAC: Right ventricular fractional area change
SV: Stroke volume
SVi: Stroke volume index
TAPSE: Tricuspid annular plane systolic excursion
TPAVF: Time of peak aortic valve flow velocity

## Author contributions

All of the authors participated in the completion of this manuscript.

## Acknowledgments

We acknowledge the financial support from the National Natural Science Foundation of China (grant 81922033 to Li Zhang; grant 81727805 to MX; and grant 81701716 to ZS) and National Institute for Health Research, UK (ICA-CL-2018-04-ST2-012) to HG and by British Heart Foundation, UK (PG/19/23/34259) to HG.

